# Deep learning-based prediction of cardiopulmonary disease in retinal images of premature infants

**DOI:** 10.1101/2025.09.18.25336004

**Authors:** Praveer Singh, Sourav Kumar, Riya Tyagi, Benjamin K Young, Brian K Jordan, Brian Scottoline, Patrick D. Evers, Susan Ostmo, Aaron S Coyner, Wei-Chun Lin, Aarushi Gupta, Deniz Erdogmus, RV Paul Chan, Emily A. McCourt, James S Barry, Cindy T. McEvoy, Michael F Chiang, J Peter Campbell, Jayashree Kalpathy-Cramer

**Author notes:** Corresponding author: J Peter Campbell. co-first authors; co-Senior authors (Praveer Singh, Sourav Kumar).

## Abstract

**Importance:** Bronchopulmonary dysplasia (BPD) and pulmonary hypertension (PH) are leading causes of morbidity and mortality in premature infants.

**Objective:** To determine whether images obtained as part of retinopathy of prematurity (ROP) screening might contain features associated with BPD and PH in infants, and whether a multi-modal model integrating imaging features with demographic risk factors might outperform a model based on demographic risk alone.

**Design:** A deep learning model was used to study retinal images collected from patients enrolled in the multi-institutional Imaging and Informatics in Retinopathy of Prematurity (i-ROP) study.

**Setting:** Seven neonatal intensive care units.

**Participants:** 493 infants at risk for ROP undergoing routine ROP screening examinations from 2012 to 2020. Images were limited to <=34 weeks post-menstrual age (PMA) so as to precede the clinical diagnosis of BPD or PH.

**Exposure:** BPD was diagnosed by the presence of an oxygen requirement at 36 weeks PMA, and PH was diagnosed by echocardiogram at 34 weeks. A support vector machine model was trained to predict BPD, or PH, diagnosis using: A) image features alone (extracted using Resnet18), B) demographics alone, C) image features concatenated with demographics. To reduce the possibility of confounding with ROP, secondary models were trained using only images without clinical signs of ROP.

**Main Outcome Measure:** For both BPD and PH, we report performance on a held-out testset (99 patients from the BPD cohort and 37 patients from the PH cohort), assessed by the area under receiver operating characteristic curve.

**Results:** For BPD, the diagnostic accuracy of a multimodal model was 0.82 (95% CI: 0.72-0.90), compared to demographics 0.72 (0.60-0.82; P=0.07) or imaging 0.72 (0.61-0.82; P=0.002) alone. For PH, it was 0.91 (0.71-1.0) combined compared to 0.68 (0.43-0.9; P=0.04) for demographics and 0.91 (0.78-1.0; P=0.4) for imaging alone. These associations remained even when models were trained on the subset of images without any clinical signs of ROP.

**Conclusions and Relevance:** Retinal images obtained during ROP screening can be used to predict the diagnosis of BPD and PH in preterm infants, which may lead to earlier diagnosis and avoid the need for invasive diagnostic testing in the future.

**KEY POINTS:** *Question:* Can an artificial intelligence (AI) algorithm diagnose bronchopulmonary dysplasia (BPD) or pulmonary hypertension (PH) in retinal images in preterm infants obtained during retinopathy of prematurity (ROP) screening examinations?

*Findings:* AI was able to predict the presence of both BPD and PH in retinal images with higher accuracy than what could be predicted based on baseline demographic risk alone.

*Meaning:* Deploying AI models using images obtained during retinopathy of prematurity screening could lead to earlier diagnosis and avoid the need for more invasive diagnostic testing.

## INTRODUCTION

Bronchopulmonary dysplasia (BPD) is a leading cause of morbidity in premature infants, affecting one-third of very preterm infants, or ∼18,000 babies annually in the United States^1^. Not only does BPD result in longer hospitalization with a higher risk of prolonged mechanical ventilation and oxygen exposure, but it also is associated with substantial long-term pulmonary and neurodevelopmental morbidity^2,3^. Pulmonary hypertension (PH) is a separate, but related condition characterized by elevation in pulmonary vascular pressure^4–6^ PH can also occur in infants with BPD (BPD-related PH), which adds significant complexity to clinical care, with 50% mortality within the first 2 years if there is both severe BPD with severe PH, enhanced risk for tracheostomy, and a 4.6x risk of mortality compared to BPD infants without PH^7,8^.

Clinical diagnosis of PH and prediction of BPD can be challenging. The most commonly used BPD prediction tools^9–11^ rely primarily on demographic data such as gestational age at birth, birth weight, race/ethnicity, and oxygen exposure, which are easy to obtain but neither sensitive nor specific enough to be actionable. The gold standard for PH diagnosis is invasive cardiac catheterization, though the diagnosis is often made with echocardiography which is easier to obtain, but still resource and time intensive^12^. Obtaining alternative biomarkers correlated with earlier stages of BPD and PH through non-invasive techniques could avoid the need for more invasive testing, and potentially help neonatologists to distinguish different endotypes related to BPD as well as PH, enabling more individually tailored care.

Recent advances in deep learning (DL) have demonstrated the ability for DL algorithms to diagnose non-ocular diseases from analysis of large datasets of retinal images. Often referred to as “oculomics” ^13,14^, this emerging field has shown promise for the retinal-based diagnosis of numerous diseases including cardiovascular, neurodegenerative, renal, and neurological disorders^15–20^, among others. For an oculomics approach to be clinically valuable, fundus imaging must be easily integrated into the care pathway, and the insights gained from retinal image analysis must provide predictive and/or diagnostic value beyond what is achievable without it.

In this project, we analyzed a large dataset of fundus images obtained as part of routine retinopathy of prematurity (ROP) examinations in the neonatal intensive care unit (NICU) using DL to determine if these images might reveal patients that would later develop the clinical diagnosis of either BPD or PH. Since ROP examinations are often performed with digital fundus imaging, there would be a path to deploy predictive algorithms that demonstrate clinical value. However, because underlying demographic risk (especially gestational age [GA] at birth) is so highly correlated with ROP, BPD and PH, we wanted to evaluate whether there was added value to imaging beyond knowledge of the demographic risk, and independent of the presence of ROP.

## METHODS

### Datasets

The study dataset was developed as part of a multi-institutional Imaging and Informatics in Retinopathy of Prematurity (i-ROP) study^21^ between January 2012 and July 2020. All infants hospitalized at participating clinical centers were eligible for recruitment if they met published criteria for requiring ROP screening exams with <1501 gm birthweight (BW) or < 31-week GA in the NICU. Patients were excluded if they had other structural ocular anomalies or if they were considered unstable for examination by their attending neonatologist. For each infant, in addition to their GA, BW, and post-menstrual age (PMA), five field-of-view images were captured using the Retcam camera system (Natus Medical Inc, Middleton, WI, USA): posterior (i.e. central), nasal, temporal, inferior, and superior. For this study, only posterior photos including the macula and optic nerve were used. As part of the original study, each eye exam was also graded for signs of ROP using the International Classification of ROP (ICROP3) criteria^22^. To reduce the potential for confounding by indication, we categorized the images into two categories: with or without signs of visible ROP.

The study was approved by the Institutional Review Boards at the coordinating center (Oregon Health & Science University) and each of the seven study centers (Columbia University, University of Illinois at Chicago, William Beaumont Hospital, Children’s Hospital Los Angeles, Cedars-Sinai Medical Center, University of Miami, Weill Cornell Medical Center) and was conducted in accordance with the Declaration of Helsinki^23^. Informed consent was obtained from parents of all infants enrolled in this study.

BPD was defined as neonates who required oxygen support at 36 weeks PMA^24^. Using this definition, we identified babies that had been diagnosed with BPD from the entire i-ROP cohort sites (BPD dataset), comprising a total of 1357 deidentified posterior pole Retinal fundus Photos (RFPs) from 493. Full inclusion/exclusion criteria have been outlined in the supplemental figure and table.

The PH dataset was a subset of the full i-ROP dataset from a single site (Oregon Health & Science University) where additional echocardiographic categorical labels for PH were available. PH was defined based on an echocardiogram at 34 weeks PMA and recorded as a binary variable defined as pulmonary artery pressure greater than ½ systemic pressure as estimated by echocardiogram^25^. This echocardiographic determination was made by the interpreting cardiologist using tricuspid valve regurgitation jet velocity, post-tricuspid shunt velocity, end-systolic interventricular septal position, or pulmonary valve regurgitation jet velocity. The PH retinal image dataset comprised 616 de-identified posterior pole RFPs from 184 patients. Full inclusion/exclusion criteria have been outlined in the supplemental figure and table.

### Algorithm Development

#### Bronchopulmonary Dysplasia

We first divided the 493 patients into 80% for Training (394 patients) and 20% for a held-out Test set (99 patients). The training set was further divided into training and validation splits at the patient level. We first trained our image feature extractor deep learning (DL) model (Resnet18 architecture^26^ as shown in **Figure 1**) to predict subsequent BPD diagnosis using RFPs labeled as BPD or no BPD as inputs to the model. We ran the model training for 30 epochs using a batch size of 64, Adam optimizer with an initial learning rate of 1e-4 and used Binary Cross Entropy as a loss function. Owing to the unequal proportion of Normal vs BPD samples, we used equal-weighted sampling for our model training. Input images were resized to 480×640 and processed with a CLAHE normalization^27^. We used standard data augmentations, including random flip (50%), random rotation, color jitter, gaussian blur and random affine. Finally, we used early stopping, choosing the best-performing model with the highest Area under the Receiver Operating Curve (AUC-ROC) score on the validation set. To avoid any confounding with ROP, a secondary model was trained on images without visible signs of ROP. Both models were then evaluated on a held-out test set (described in the evaluation section). For the multi-modality model, as shown in **Figure 1**, the RFPs were first passed as input into the Resnet18 feature extraction model, resulting in a 512-dimensional feature vector per image (before the final fully connected layer and after the last average pooling layer). This 512-dimensional feature vector is then concatenated with BW, GA, and PMA to train a shallow Support Vector Machine (SVM)^28^ classifier model using the integrated features to predict BPD at 36 weeks PMA. The SVM was trained using a Radial Basis Function (RBF) kernel with a default regularization parameter (1) and kernel coefficient (1/size of extracted feature vector).

**Figure 1:**
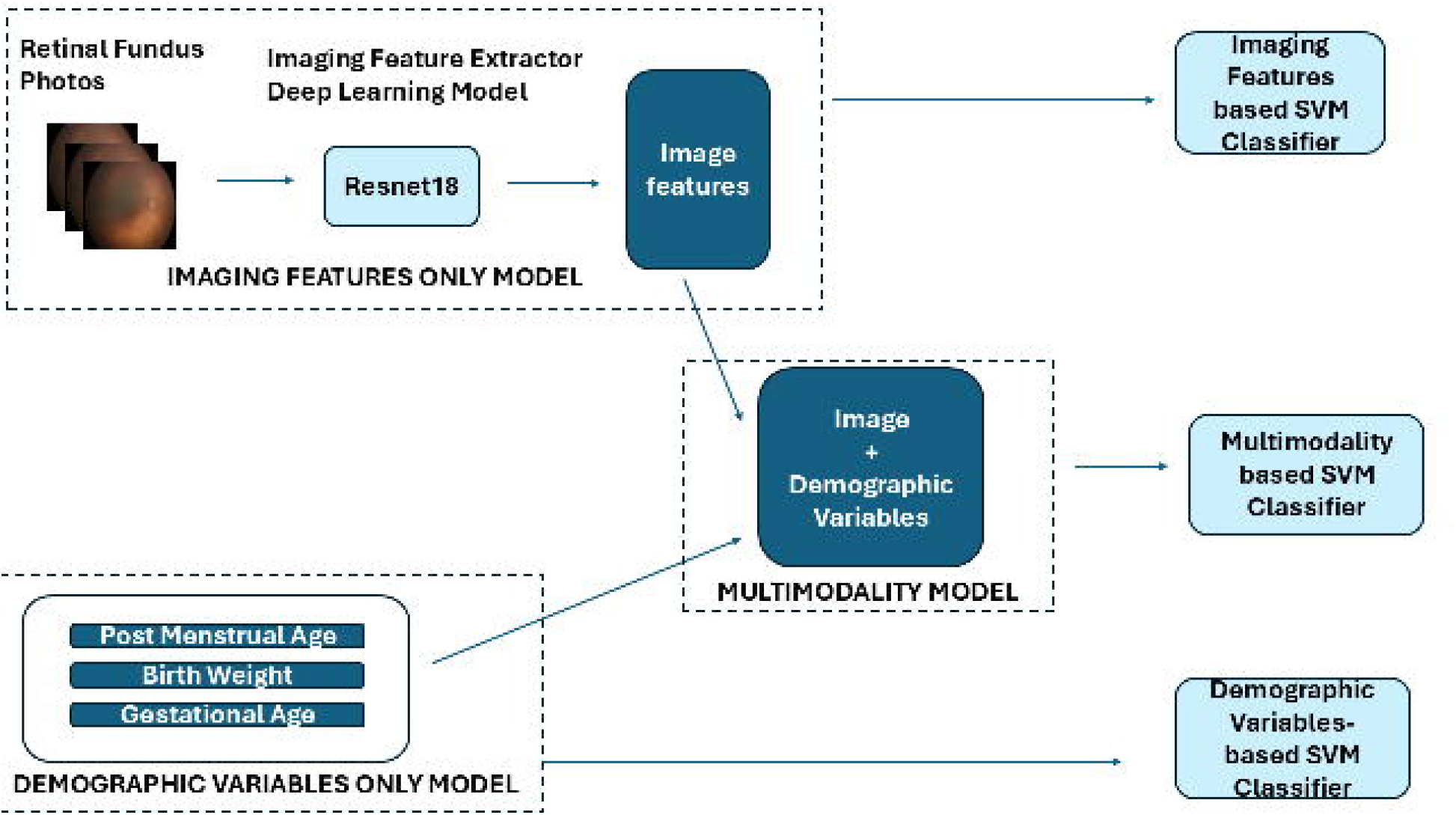
Workflow of Imaging, Demographic, and Multimodal Models for BPD and PH Classification. Modeling framework for classifying neonatal cardiopulmonary outcomes using retinal fundus images and demographic variables. The Imaging Features Only Model uses retinal fundus photographs processed through a deep learning feature extractor (ResNet18) to generate image features, which are then classified using a Support Vector Machine (SVM). The Demographic Variables Only Model utilizes postmenstrual age, birth weight, and gestational age as inputs to an SVM classifier. The Multimodality Model combines both image features and demographic variables, which are jointly inputed into a multimodal SVM classifier for improved predictive performance. The three models operate independently for performance comparison and evaluation.

#### Pulmonary Hypertension

Similar to the BPD cohort, we divided the 184 patients in the PH cohort into 80:20 splits for training (147 patients) and testing (37 patients) sets. Likewise, the training set was further divided into training and validation (on the patient level). Due to the limitation of the size of the dataset, we could not fine-tune the Resnet18 model and instead extracted image features using an Imagenet pre-trained version of Resnet18. We finally trained a shallow SVM classifier model over the extracted features to predict PH at 34 weeks PMA. For the multi-modality model, similar to BPD, we used the concatenated RFP features and clinical variables to predict PH. Again like BPD, each SVM for PH was trained using an RBF kernel with a default regularization parameter (1) and kernel coefficient (1/size of extracted feature vector). Similar to BPD, we again trained a second model on images without signs of ROP.

### Evaluation and statistics

For both BPD and PH, we evaluated the performance of all models trained with different input combinations (image features alone, GA alone, integrated image features and GA) on the held out test set. We computed the AUROC score for all models as well as their 95% confidence interval range for models trained on the full dataset, and models trained only on the subset without signs of ROP. A DeLong test at a significance level of p = 0.05 was used for statistical comparisons of model performance.

## RESULTS

### Demographics

Table 1. describes the demographics for both the BPD and PH populations, including the subset of patients without evidence of ROP in the fundus images. Not surprisingly, babies with both BPD and PH had lower BW and GA, demonstrating the importance of controlling for these variables in predictive models. Full dataset demographic statistics can be found in Supplementary **Table 1**.

**Table 1:** Train Dataset characteristics for Bronchopulmonary Dysplasia and Pulmonary Hypertension for with and without ROP images.

| Metric | With ROP |  | Without ROP |  |
| --- | --- | --- | --- | --- |
|  | BPD (n=129) | Normal (n=186) | BPD (n=125) | Normal (n=183) |
| **Birthweight ± SD** | 770.18 ± 242.71 | 999.36 ± 254.31 | 781.48 ± 247.35 | 1008.70 ± 248.78 |
| **Gestational Age ± SD** | 25.59 ± 1.60 | 26.94 ± 1.79 | 25.72 ± 1.59 | 27.01 ± 1.74 |
| **Postmenstrual Age ± SD** | 32.74 ± 0.88 | 32.75 ± 0.89 | 32.69 ± 0.88 | 32.74 ± 0.89 |
| **Number of Images (%)** | 374 (42.65) | 503 (57.35) | 343 (41.08) | 492 (58.92) |
| **Number of Subjects (%)** | 129 (40.95) | 186 (59.05) | 125 (40.58) | 183 (59.42) |
|  | PH (n=14) | Normal (n=103) | PH (n=14) | Normal (n=102) |
| **Birthweight ± SD** | 716.69 ± 145.07 | 925.34 ± 280.33 | 725.44 ± 144.38 | 933.69 ± 280.55 |
| **Gestational Age ± SD** | 24.96 ± 1.44 | 26.63 ± 1.70 | 24.99 ± 1.36 | 26.70 ± 1.68 |
| **Postmenstrual Age ± SD** | 32.43 ± 0.98 | 32.54 ± 1.00 | 32.42 ± 0.98 | 32.51 ± 0.99 |
| **Number of Images (%)** | 61 (15.93) | 322 (84.07) | 36 (10.34) | 312 (89.66) |
| **Number of Subjects (%)** | 14 (11.97) | 103 (88.03) | 14 (12.07) | 102 (87.93) |

### Prediction of BPD

Figure 2. summarizes the performance of models using demographic variables alone, imaging features alone, and in combination in both the full training dataset, and the subset without ROP with 95% confidence intervals. Clinical variables alone (BW, GA and PMA) had an AUROC of 0.72 [0.73 in the non-ROP training subset]. The imaging alone model performed similarly, 0.72 [0.73]. In combination, when clinical demographics were concatenated with imaging features, the AUROC improved to 0.82 [0.78] (p<0.05 compared to imaging alone and p=0.07 compared to demographics alone). A full summary of comparative analysis between imaging and clinical-informatics input variables for predicting BPD can be found in Supplemental **Table 2 and 3**.

**Figure 2:**
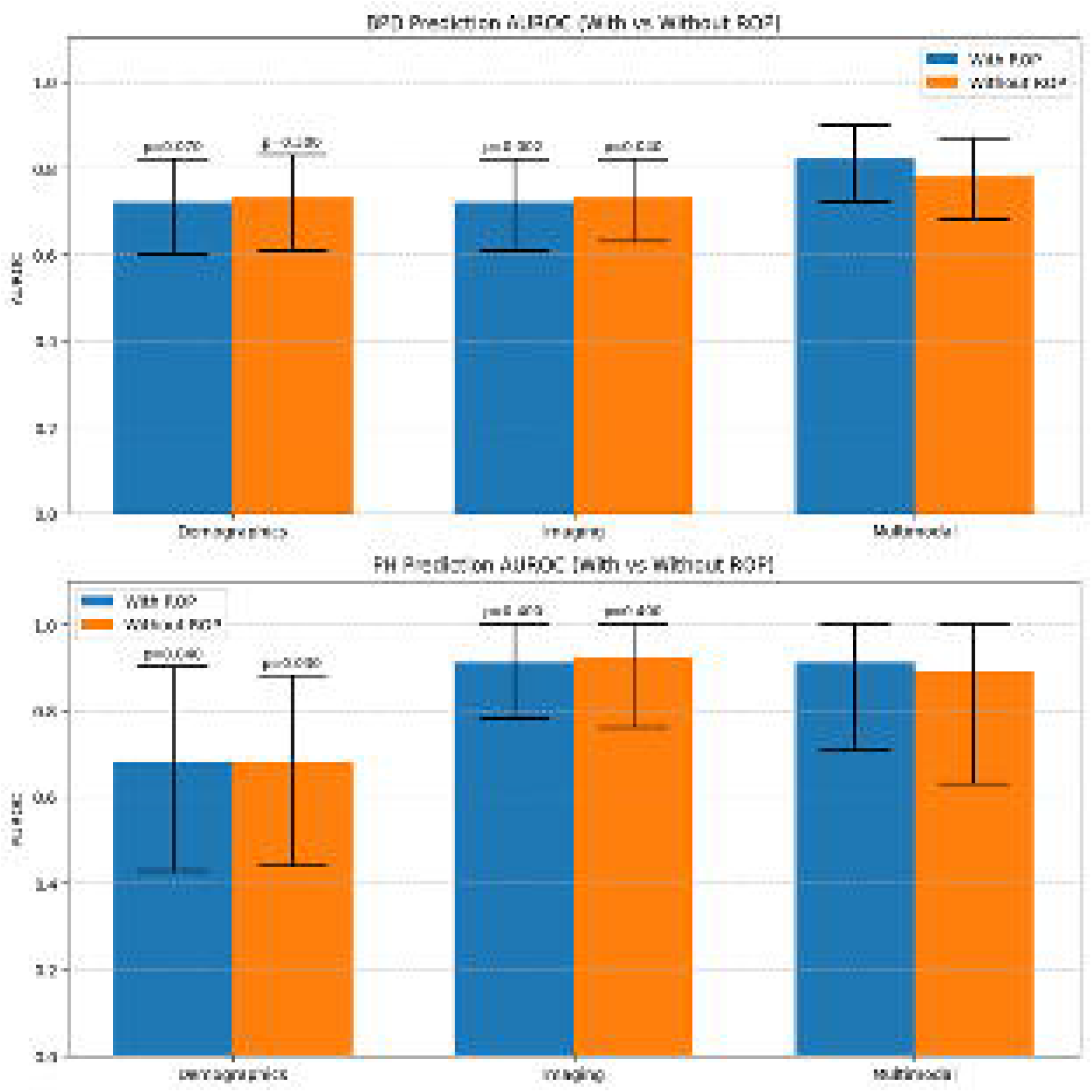
AUROC Comparisons for BPD and PH Prediction Models in Infants With and Without ROP. Area Under the Receiver Operating Characteristic Curve (AUROC) performance for predicting Bronchopulmonary Dysplasia (BPD) (top panel) and Pulmonary Hypertension (PH) (bottom panel) using three different model types: Demographics-only, Imaging-only, and Multimodal, stratified by presence or absence of Retinopathy of Prematurity (ROP). Blue bars represent infants with ROP; orange bars represent those without ROP. For each outcome, p-values indicate the statistical significance of the performance difference between the respective model and the multimodal model within each subgroup (with or without ROP). For example, in PH prediction, the multimodal model shows significantly improved performance compared to the demographics-only model (p = 0.040 for ROP; p = 0.030 for non-ROP), while no significant differences were observed with the imaging-only model. Error bars indicate 95% Confidence Interval for model performance.

**Figure 3:**
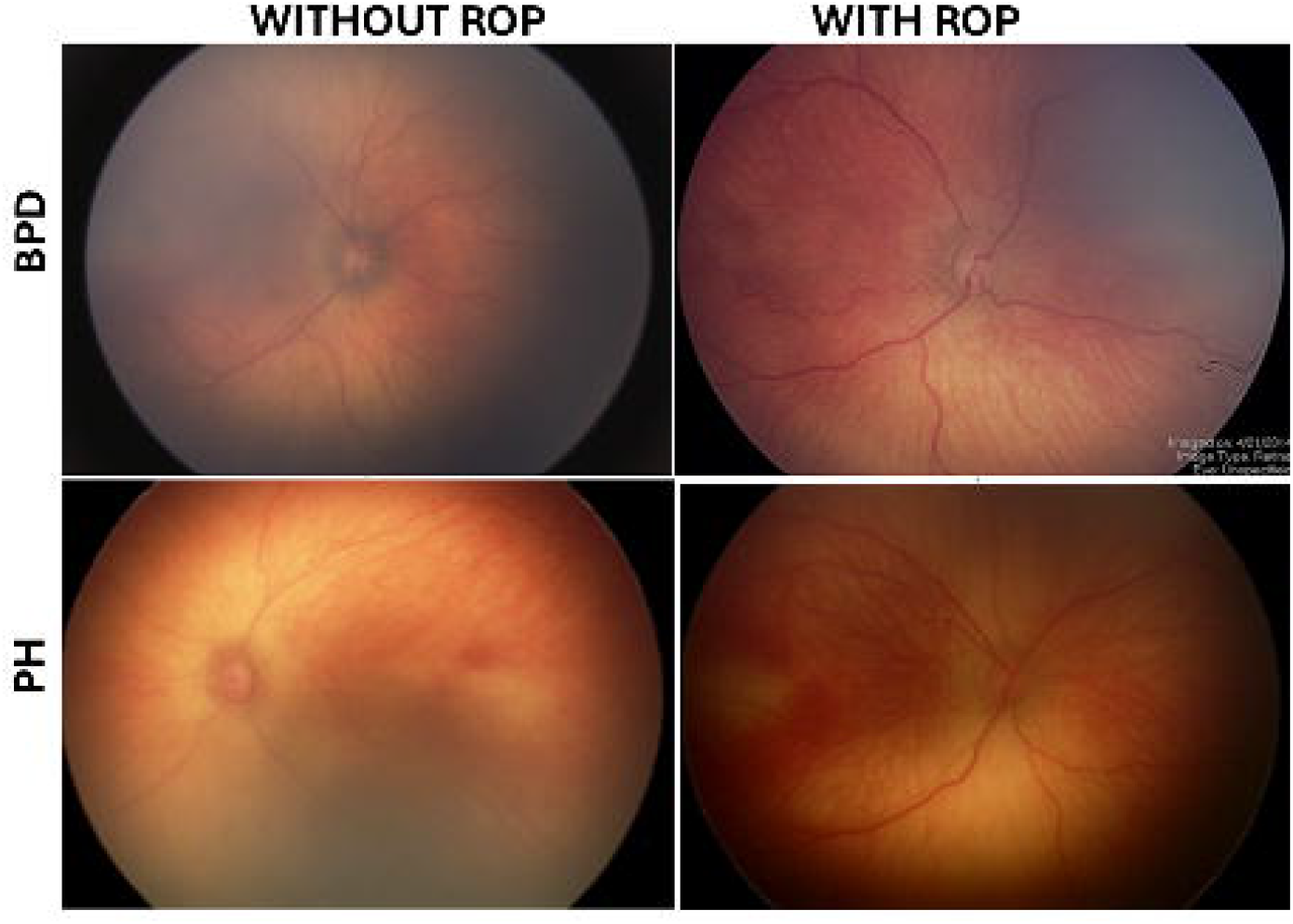
Representative Retinal Fundus Images of Infants with and Without ROP Across BPD and PH Cohorts. Retinal fundus photographs from infants diagnosed with Bronchopulmonary Dysplasia (BPD) (top row) and Pulmonary Hypertension (PH) (bottom row), stratified by the presence or absence of Retinopathy of Prematurity (ROP). Images in the left column correspond to infants *without ROP*, while those in the right column correspond to infants *with ROP*. No significant difference was observed in the predictive performance of BPD and PH models when trained with and without visible ROP as shown in Supplementary **Table 2**.

### Prediction of PH

Figure 2. summarizes the performance of models using demographic variables alone, imaging features alone, and in combination in both the full training dataset, and the subset without ROP with 95% confidence intervals. Clinical features alone (BW, GA and PMA) had an AUROC of 0.68 [0.68 in the non-ROP training subset]. The imaging alone model performed quite strongly with an AUROC 0.91 [0.92] (p<0.04 compared to demographics alone). In combination, when clinical demographics were concatenated with imaging features, there was no further improvement (0.91 [0.89]). A full summary of comparative analysis between imaging and clinical-informatics input variables for predicting BPD can be found in Supplemental **Table 2 and 3**.

## DISCUSSION

These results suggest that there is information present in retinal images that can identify the presence of cardiopulmonary disease in babies born less than 31 weeks GA and 1501grams BW, expanding the field of oculomics into the pediatric population. In the case of BPD, the images appeared to complement the “pretest” risk based on demographics alone. In the case of PH, the images were highly diagnostic independent of demographic information. Neither of these results appeared to be due to confounding by indication of the underlying diagnosis of ROP, but related to non-ROP imaging features.

In the case of BPD, there appears to be added information in RFPs collected before 34 weeks PMA that is associated with an ongoing oxygen requirement at 36 weeks (the classic definition of BPD), which is more prognostic than simply knowledge of underlying risk based on BW and GA. The pathophysiology of BPD is multifactorial. It may be that various pulmonary phenotypes have different ocular manifestations, which could lead to earlier recognition and more targeted interventions for various endotypes of BPD. It is likely that babies who remain on oxygen at 36 weeks have “ocular manifestations” for higher oxygen requirements for both oxygen concentration and/or continuous positive airway pressure (CPAP) in earlier time points, and it may be that the features “learned” by the model relate to this. For example, it is plausible that mechanical ventilation of CPAP leads to changes in the retinal or choroidal vasculature that the algorithm is able to learn. In this dataset, we did not have access to the oxygen dose and delivery mechanism at every time point, but future work could evaluate whether the imaging findings are reflective only of oxygen exposure (which can be determined without imaging) or independently predictive.

For PH, the retinal images were highly diagnostic. It is likely that elevated right heart pressures lead to higher jugular venous pressure, delayed retinal venous drainage, and changes in the retinal or choroidal vasculature, as has been described in adults ^29–31^. Current practice for PH screening in high risk babies involves the use of echocardiography, which is a limited resource and requires specialized expertise both for image acquisition and interpretation, and at some institutions standard procedure involves invasive cardiac catheterization for confirmation of severe BPD PH. If further work suggests that the retinal images could identify PH prior to standard screening, then it may provide a window for earlier testing and management, as well as potentially spare a higher risk catheterization procedure where that remains standard of care.

There have been several oculomics applications that lack practical or clinical value. Some, such as the prediction of biologic sex^32^ or smoking status^15,16^, have alternative methods of assessment and do not require retinal imaging. Others may not have added value over a thorough history, physical exam, or easily obtainable laboratory evaluation, or be neither sensitive nor specific enough to be actionable in any meaningful way. Finally, since fundus photography is not widely performed outside of eye care practices, in other cases, even if there is added value, there is no practical method to deploy the technology. However, the methods described herein differ in several important ways. All babies born < 31 weeks GA or < 1500 grams require ROP screening, which is increasingly performed using fundus photography, providing a built-in deployment mechanism for these types of algorithms, should they prove useful. One of the strengths of this analysis is that we compared the diagnostic performance of the algorithm with the underlying demographic risk, and both in the case of BPD and especially in the case of PH, the images appeared to add value. In practice, deployment of any diagnostic test will occur in a particular clinical context with a known pretest probability. In our case, we combined the diagnostic information of both demographic risk and imaging features into a single model by concatenating the demographic information into the feature vector from the ResNet output, but there would be several other potential ways to combine this information. The results suggest that combining imaging data with clinical data might yield higher diagnostic predictive capability than either modality alone.

There are several limitations to this analysis. First, we anticipate that because of the common shared risk factors for all of these outcomes, it is possible that some of the features learned relate simply to the presence of eye disease, and that babies with more severe eye disease are more likely to have BPD or PH (for e.g. past works have demonstrated that one of the clinical signs of PH can be increased dilation/tortuosity of the vessels which is linked to severity of ROP disease^29,31^). We attempted to control this by removing retinal images with visible ROP, including the presence of pre-plus or plus disease, and found results to be consistent across both experiments. Second, deep learning models tend to be brittle and perform poorly when tested on out-of-distribution datasets, which we did not do in this study^33^. Third, we duly acknowledge that the power of this study limits the statistical significance of some of our results. Fourth, we are of course curious about what features the AI models are learning, though it is not always easy to answer this, and we did not include any explainability analyses, in part because many of the common post-hoc saliency map techniques fail basic validation testing, such as repeatability and robustness^34^. Nonetheless, exploring these features with validated explainability measures will be an important future step. Lastly, we did not directly compare our model to all the reported clinical models and tools for prediction of BPD and PH, only to the risk of BPD and PH based on the degree of prematurity alone. It may be that there are clinically available models including other factors (such as oxygen exposure) that work as well as the models reported here, which would reduce the significance of these findings.

In conclusion, we showcase the first proof-of-concept oculomics investigation in neonates. We demonstrate that retinal fundus images obtained as early as 31-32 weeks PMA can identify a future diagnosis of BPD. We further show that there is information in the images relevant to the diagnosis of PH, which requires echocardiography, and in some instances, invasive testing to confirm. Finally, we demonstrate that a combined imaging and clinical informatics multimodality AI model improves performance for the prediction of BPD and PH in comparison to an image-only or demographics-only model, respectively. We consider these results to be hypothesis-generating and should be more fully evaluated in larger, heterogeneous populations and in images from other camera systems and modalities. Nonetheless, the potential for earlier, non-invasive methods of detecting comorbidities of prematurity may open new doors of clinical management and scientific inquiry, which may lead to improved outcomes in this challenging population.

## Supporting information

ACCEPT - AI Guidelines

TRIPOD AI Checklist

## Data Availability

All code for model training and analysis can be found at the following github link: https://github.com/QTIM-Lab/AI-for-BPD-and-PH.git. Patient data cannot be made available due to privacy concerns.

## Funding

This work was supported by grants R01 EY19474, R01 EY031331, R21 EY031883, and P30 EY10572 from the National Institutes of Health (Bethesda, MD) and by unrestricted departmental funding and a Career Development Award (JPC) from Research to Prevent Blindness (New York, NY). Additionally, this research was supported in part by the NIH Intramural Research Program of the National Institutes of Health (NIH). The contributions of the NIH author were made as part of their official duties as NIH federal employees, are in compliance with agency policy requirements, and are considered Works of the United States Government. However, the findings and conclusions presented in this paper are those of the authors and do not necessarily reflect the views of the NIH or the U.S. Department of Health and Human Services.

## Conflict of Interest Disclosures

- Drs. Campbell, Chan, and Kalpathy-Cramer receive research support from Genentech (San Francisco, CA). Dr. Chiang previously received research support from Genentech.
- Dr. Chan is a consultant for Alcon (Ft Worth, TX).
- Dr. Barry is an AI and technology forum lead for Healio.
- Drs. Chan, Kalpathy-Cramer, Coyner, and Campbell are equity owners of Siloam Vision.

## Notes

### Clinical Protocols

https://clinicaltrials.gov/study/NCT04420156?id=%23%20NCT04420156&rank=1

### Author Declarations

The study was approved by the Institutional Review Boards at the coordinating center (Oregon Health & Science University) and each of the seven study centers (Columbia University University of Illinois at Chicago William Beaumont Hospital Childrens Hospital Los Angeles Cedars-Sinai Medical Center University of Miami Weill Cornell Medical Center) and was conducted in accordance with the Declaration of Helsinki23. Informed consent was obtained from parents of all infants enrolled in this study.

