## Supplementary 1 for "Deep learning-based prediction of cardiopulmonary disease in retinal images of premature infants"

**SUPPLEMENTARY MATERIAL**

**Data and Code**

All code for model training and analysis can be found at the following github link: <https://github.com/QTIM-Lab/AI-for-BPD-and-PH.git>. Patient data cannot be made available due to privacy concerns.

**ACCEPT-AI reporting guidelines**

A duly completed ACCEPT-AI reporting guidelines checklist (<https://www.nature.com/articles/s41746-023-00898-5>) can be found in the supplementary material.

**TRIPOD-AI reporting guidelines**

A duly completed TRIPOD-AI reporting guidelines checklist (<https://www.bmj.com/content/385/bmj-2023-078378>) can be found in the supplementary material.

**Split Creation Heuristic**

While creating the training, testing, and validation splits, we ensured similar distributions both across disease labels as well as the clinical variable values. This is challenging as the labels are discrete, while the clinical variables are continuous values. To create splits that have similar distribution of labels and all clinical variables, we treat each of the clinical variables (BW, PMA and GA) as discrete variables by binning them into quartile ranges. Stratified splits were then created ensuring nearly equal frequency of all possible combinations of disease labels and discrete clinical variable values across the Train/Test/Val splits.

**Figure 1: Study profile of BPD and PH cohorts.**

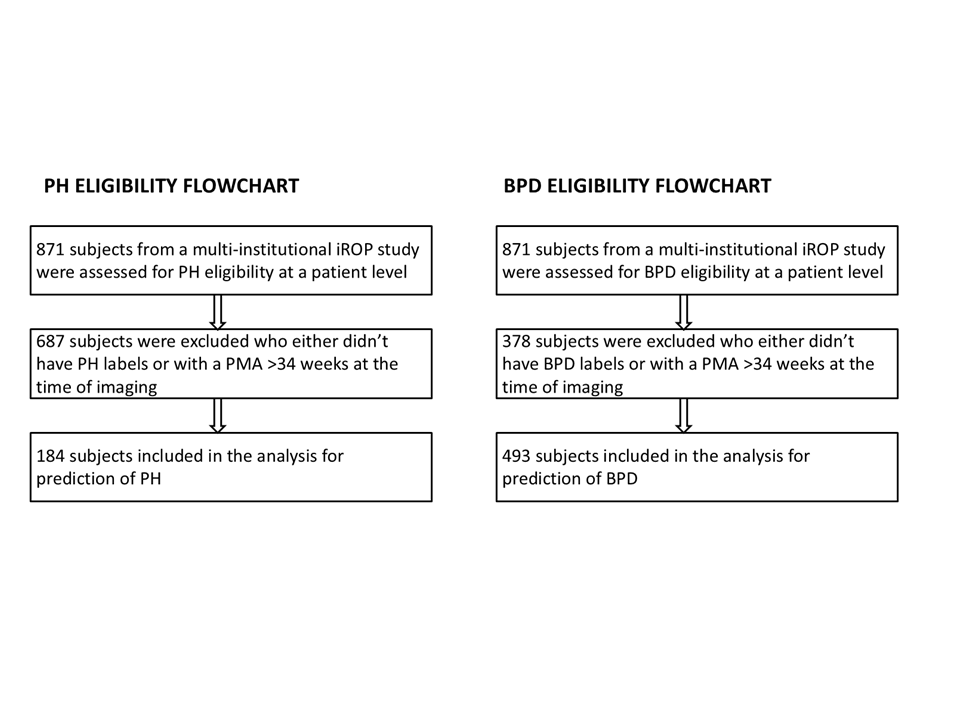

A total of 871 subjects from a multi-institutional iROP study were assessed for eligibility. For bronchopulmonary dysplasia (BPD) analysis (left panel), 378 subjects were excluded due to either missing BPD labels or postmenstrual age (PMA) greater than 34 weeks at the time of imaging, resulting in 493 subjects included in the final analysis. For pulmonary hypertension (PH) analysis (right panel), 687 subjects were excluded due to either missing PH labels or PMA greater than 34 weeks, resulting in 184 subjects included in the final analysis.

**Table 1: Full Dataset characteristics for Bronchopulmonary Dysplasia and Pulmonary Hypertension.**

| **Metric** | **BPD** (n=202) | **Normal** (n=291) | **PH** (n=23) | **Normal** (n=161) |
| --- | --- | --- | --- | --- |
| **Birthweight ± SD** | 805.86 ±252.35 | 1035.72 ±274.15 | 718.4 ± 183.1 | 920.2 ± 270.6 |
| **Gestational Age ± SD** | 25.72 ±1.75 | 27.26 ±1.81 | 25.11 ± 1.534 | 26.62 ± 1.615 |
| **Postmenstrual Age ± SD** | 32.39 ±0.84 | 32.54 ±0.95 | 32.43 ± 1.010 | 32.59 ± 0.9917 |
| **Number of Images (%)** | 575 (42.37) | 782 (57.63) | 100 (16.23) | 516 (83.77) |
| **Number of Subjects (%)** | 202 (40.97) | 291 (59.03) | 23 (12.50) | 161 (87.50) |

**Table 2: Comparative analysis of with and without ROP for the prediction of BPD and PH.**

| **Input Variables** | **AUROC for BPD (95% C.I.)**  **(with ROP)** | **AUROC for BPD (95% C.I.)**  **(without ROP)** | **P value** |
| --- | --- | --- | --- |
| **BW + GA + PMA (Demographics model)** | 0.72 (0.60,0.82) | 0.73 (0.61,0.83) | 0.29 |
| **Imaging** | 0.72 (0.61,0.82) | 0.73 (0.63,0.82) | 0.80 |
| **BW + GA + PMA + Imaging (Multimodal model)** | 0.82 (0.72, 0.90) | 0.78 (0.68,0.87) | 0.27 |

| **Input Variables** | **AUROC for PH (95% C.I.)**  **(with ROP)** | **AUROC for PH (95% C.I.)**  **(without ROP)** | **P value** |
| --- | --- | --- | --- |
| **BW + GA + PMA (Demographics model)** | 0.68 (0.43, 0.9) | 0.68 (0.44,0.88) | 0.81 |
| **Imaging** | 0.91 (0.78,1.0) | 0.92 (0.76,1.0) | 0.79 |
| **BW + GA + PMA + Imaging (Multimodal model)** | 0.91 (0.71,1.0) | 0.89 (0.63,1.0) | 0.91 |

**Table 3: Predictive accuracy of various imaging and clinical-informatics input variables for prediction of BPD and PH, both with and without ROP. P-values are shown in comparison to the multimodality model.**

| **Input Variables** | **AUROC for BPD (95% C.I.)**  **(with ROP)** | **AUROC for BPD (95% C.I.)**  **(without ROP)** | **P value (with ROP, without ROP) v/s Multimodal Model** |
| --- | --- | --- | --- |
| **BW** | 0.70 (0.57,0.80) | 0.69 (0.57,0.79) | 0.03,0.1 |
| **GA** | 0.73 (0.61,0.83) | 0.73 (0.62,0.83) | 0.06,0.35 |
| **PMA** | 0.51 (0.40,0.63) | 0.53 (0.41,0.64) | 0.0005,0.0007 |
| **BW + GA + PMA (Demographics model)** | 0.72 (0.60,0.82) | 0.73 (0.61,0.83) | 0.07,0.34 |
| **Imaging** | 0.72 (0.61,0.82) | 0.73 (0.63,0.82) | 0.002,0.042 |
| **BW + Imaging** | 0.79 (0.70,0.88) | 0.76 (0.67,0.85) | 0.1,0.2 |
| **GA + Imaging** | 0.78 (0.68,0.87) | 0.76 (0.66,0.85) | 0.04,0.09 |
| **PMA + Imaging** | 0.72 (0.61,0.82) | 0.73 (0.63,0.82) | 0.03,0.03 |
| **BW + GA + PMA + Imaging (Multimodal model)** | 0.82 (0.72, 0.90) | 0.78 (0.68,0.87) |  |

| **Input Variables** | **AUROC for PH (95% C.I.)**  **(with ROP)** | **AUROC for PH**  **(95% C.I.)**  **(without ROP)** | **P value (with ROP, without ROP) v/s Multimodal Model** |
| --- | --- | --- | --- |
| **BW** | 0.67 (0.47, 0.87) | 0.65 (0.45,0.86) | 0.02,0.03 |
| **GA** | 0.6 (0.18, 0.96) | 0.77 (0.58,0.93) | 0.09,0.12 |
| **PMA** | 0.52 (0.23, 0.86) | 0.58 (0.26,0.83) | 0.02,0.1 |
| **BW + GA + PMA (Demographics model)** | 0.68 (0.43, 0.9) | 0.68 (0.44,0.88) | 0.04,0.03 |
| **Imaging** | 0.91 (0.78,1.0) | 0.92 (0.76,1.0) | 0.4,0.46 |
| **BW + Imaging** | 0.89 (0.78,1.0) | 0.92 (0.72,1.0) | 0.6,0.36 |
| **GA + Imaging** | 0.88 (0.70,1.0) | 0.88 (0.61,1.0) | 0.7,0.47 |
| **PMA + Imaging** | 0.9 (0.77,1.0) | 0.92 (0.76,1.0) | 0.6,0.46 |
| **BW + GA + PMA + Imaging (Multimodal model)** | 0.91 (0.71,1.0) | 0.89 (0.63,1.0) |  |

**Table 4: Subject Inclusion Criteria for Bronchopulmonary Dysplasia (top) and Pulmonary Hypertension (bottom)**

| **Inclusion Criteria** | **Determined by** |
| --- | --- |
| Birthweight < 1501 | Examining Physician |
| Gestational Age < 31 weeks | Examining Physician |
| PMA <=34 weeks | Examining Physician |
| BPD (on oxygen support at 36 weeks PMA) | Neonatologist |
| Negative for plus disease diagnosis (for ROP-Constrained experiment) | ROP Expert Consensus |

| **Inclusion Criteria** | **Determined by** |
| --- | --- |
| Birthweight < 1501 | Examining Physician |
| Gestational Age < 31 weeks | Examining Physician |
| PMA <=34 weeks | Examining Physician |
| Detected with PH at 34 weeks PMA | Cardiologist through echocardiogram |
| Negative for plus disease diagnosis (for ROP-Constrained experiment) | ROP Expert Consensus |

**Table 5: Race Distribution for Bronchopulmonary Dysplasia (top) and Pulmonary Hypertension (bottom)**

| **Race** | **Count (%)** |
| --- | --- |
| Caucasian/White | 332 (67.3%) |
| African American | 95 (19.3%) |
| Chinese | 11 (2.2%) |
| Middle Eastern | 8 (1.6%) |
| Korean | 5 (1.0%) |
| Filipino | 5 (1.0%) |
| Asian Indian | 4 (0.8%) |
| Other Asian -> Specify | 3 (0.6%) |
| American Indian or Alaska Native | 2 (0.4%) |
| Japanese | 2 (0.4%) |
| Other Native Hawaiian or Pacific Islander -> Specify | 1 (0.2%) |
| Vietnamese | 1 (0.2%) |
| Missing/Unreported | 24 (4.9%) |

| **Race** | **Count (%)** |
| --- | --- |
| Caucasian/White | 163 (88.6%) |
| African American | 13 (7.1%) |
| Filipino | 3 (1.6%) |
| Vietnamese | 2 (1.1%) |
| American Indian or Alaska Native | 1 (0.5%) |
| Missing/Unreported | 2 (1.1%) |

**Table 6: Ethnicity Distribution for Bronchopulmonary Dysplasia (top) and Pulmonary Hypertension (bottom)**

| **Ethnicity** | **Count (%)** |
| --- | --- |
| Other Hispanic -> Specify | 52 (10.6%) |
| Mexican | 47 (9.5%) |
| Cuban | 30 (6.1%) |
| Dominican | 20 (4.1%) |
| Hispanic, Unknown Type | 4 (0.8%) |
| Puerto Rican | 4 (0.8%) |
| Missing/Unreported | 336 (68.1%) |

| Ethnicity | Count (%) |
| --- | --- |
| Mexican | 35 (19.0%) |
| Hispanic, Unknown Type | 2 (1.1%) |
| Other Hispanic -> Specify | 3 (1.6%) |
| Missing/Unreported | 144 (78.3%) |

**Table 7: Sex Distribution for Bronchopulmonary Dysplasia (top) and Pulmonary Hypertension (bottom)**

| **Sex** | **Count (%)** |
| --- | --- |
| Male | 267 (54.2%) |
| Female | 226 (45.8%) |

| Sex | Count (%) |
| --- | --- |
| Male | 106 (57.6%) |
| Female | 78 (42.4%) |
