## Supplementary material for "Deep learning-based prediction of cardiopulmonary disease in retinal images of premature infants": ACCEPT - AI Guidelines

ACCEPT-AI Reporting Checklist:

1. Age

• Include the patient’s chronological age at the time of study enrollment. -Reported the mean Gestational Age and Postmenstrual age of the study population.

• Include the patient’s developmental age when applicable and available; if unavailable, state this. - N/A

• Attempt to include developmental stages and relevant milestone metrics of CYP (e.g., height and weight percentile upon enrollment) to capture participant heterogeneity; if developmental metrics are unavailable, state this. - N/A

• Include the age(s) of intended algorithm users (e.g., pediatric only, pediatric and adult, or adult only). -Pediatric only

2. Communication – For all points below: This study is a post-hoc analysis of a multicenter cohort conducted approximately a decade ago. While there was no direct community or patient engagement in the original study, we did involve multiple specialty stakeholders, including ophthalmology, neonatology, and cardiology, in the research process.

• Communicate the study purpose to CYP as key stakeholders using developmentally appropriate communication strategies.

• Communicate with parent(s) or legal guardians as key stakeholders.

• Tailor communication to social circumstances, addressing family complexities, including any court involvement.

• Clearly communicate technology-specific study purpose, risks, benefits, and alternatives with all key stakeholders.

• Consider the use of videos, written material, and decision aids to facilitate education and enhance communication.

• Involve stakeholders, including CYP and parents, in focus groups for design feedback where possible and relevant legal and institutional permissions are obtained.

• State efforts taken to involve potential users in feedback of the research idea and invest in community-level digital literacy.

• Where possible, document and articulate model explainability. -Articulated in the Discussions section.

3. Consent and Assent -

• Record the mode of consent, who provided consent (e.g., parent, legal guardian), and how it was obtained. Parents provided written informed consent for the collection of data in this project.

• Document any complex parental relations, dynamics, or court involvement that impact consent. Not available.

• Document children’s social circumstances as relevant to safety, participation, and evaluation. Not available.

• For children in state custody, ensure consent is obtained by relevant legal guardians or custodians and documented accurately. N/A

• Document relevant child protection laws pertinent to individual cases. Not available.

• Attain assent when developmentally appropriate and/or required by regulations. N/A

• Ensure minors participate in the assent process in accordance with their developmental skills (e.g., appropriate modifications for children with clinically relevant developmental delay). N/A

• Record the age when assent is provided. Not available

• Ensure local laws for adolescent assent/consent are followed. N/A

4. Equity

• Ensure inclusion and exclusion criteria are clearly defined, specifying disease, symptom, or condition of interest, with developmental stages considered as appropriate. -Reported in Supplementary Figure 1 and Table 4

• State processes employed to reduce selection bias. - We systematically recruited participants from eight centers, encompassing the entire target population. As a result, the study sample is broadly representative of the intended use population. The primary source of potential selection bias lies in parental consent, which is inherently difficult to control or mitigate.

• Provide transparent demographic reporting, including race, documented sex, gender, and socioeconomic factors. - Race, ethnicity, and sex data for participants with BPD and PH are presented in Supplementary Tables 5, 6, and 7. Socioeconomic information, however, was not available for this population.

• Provide details on how gender and documented sex have been incorporated into the study design. - The sex assigned at birth was recorded for each newborn and used in the study analyses.

• Incorporate accessible research design to facilitate the inclusion of patients with disabilities (developmental and otherwise). -N/A

• If skin tone could influence algorithmic outputs, ensure it is documented. - Since this study utilizes raw fundus images containing pigmentation features for the classification of BPD and PH, there is a possibility that these features may influence algorithmic outputs. However, this potential impact has not been thoroughly investigated in the current work and remains an important avenue for future research**.**

• Indicate the source of demographic information (e.g., self-reported) as well as details on non-reporting and missingness. -Self-Reported.

• Discuss the role of community engagement in the study. - This study is a post-hoc analysis of a multicenter cohort conducted approximately a decade ago. While there was no direct community or patient engagement in the original study, we did involve multiple specialty stakeholders, including ophthalmology, neonatology, and cardiology, in the research process.

5. Protection of Data

• State how data collection aligns with study objectives. - This study is a post-hoc analysis of a multicenter cohort conducted approximately a decade ago.

• State data-sharing plans when relevant. - All models and code will be made publicly available via GitHub. However, due to privacy concerns, the associated images and clinical data will not be shared.

• State if data is identifiable or de-identified. -Data is deidentified.

• If data is de-identified, state compliance with relevant legal frameworks (e.g., HIPAA, Common Rule, GDPR). Compliance with HIPAA.

• State data protection plans, addressing unique data risks in AI/ML, including protections against cybersecurity breaches. All data are stored and analyzed within a Secure Local Compute Environment (SLCE) at the University of Colorado, which is designed to prevent cybersecurity breaches and ensure data privacy.

• Disclose whether data can or cannot be retrieved/removed in the future by parents and CYP. No retrieval/removal of patient data is allowed.

• Ensure the social context of the child (e.g., suspected or confirmed child abuse or complex social circumstance) is accounted for prior to any data releases that may involve parental requests or involvement, if available. N/A

6. Technological Considerations (Transparency of Techniques, Training, and Testing Methodology)

• Ensure algorithmic studies are tailored to the needs of the pediatric population and clearly documented in the study protocol.

• Ensure AI/ML techniques are only used when potentially beneficial to the pediatric population, and that such benefits are clearly detailed in the study protocol.

• Detail any potential harms that pediatric subjects may incur as a result of the study. While no immediate harms are evident, this is a proof-of-principle study that requires further validation before any actionable conclusions can be drawn.

• Identify measures taken to minimize risk to pediatric subjects throughout the study and post-implementation. N/A

• State measures taken to monitor and document adverse events that may affect pediatric subjects. N/A

• State outcome measures and plans to clinically evaluate algorithm performance on pediatric subjects. The outcome measure and AUROC values are provided.

• When available, utilize validated pediatric clinical scales in the clinical algorithm evaluation. N/A

• Articulate how AI/ML will be trained to recognize/account for developmental heterogeneity. N/A

• Document AI/ML methods using validated guidelines (e.g., CONSORT-AI and SPIRIT-AI). Used TRIPOD-AI

• Define data input and output (e.g., images, text) as well as the source (e.g., public dataset), and output.

• Account for age-specific factors related to disability and developmental conditions (e.g., natural disease progression) as relevant in study design, testing, and evaluation. N/A

• State if the study involves adult, pediatric, or mixed data in training and/or testing. N/A

• If the study involves both adult and pediatric data, state the purpose for this combination. N/A

• If the study involves both adult and pediatric data, state whether the same or separate algorithms were used to assess each group. N/A
